# Cannabis Use, Cognition, and Suicide Risk in Adolescents from the Adolescent Brain Cognitive Development Study

**DOI:** 10.64898/2026.11.16.26368218

**Authors:** Nadine Michel, Kate T. Tran, Natasha Reddy, J. Cobb Scott, Ruben C. Gur, Elina Visoki, Ziv Bren, Tyler M. Moore, Ran Barzilay

## Abstract

**Background:** Cannabis use has rapidly emerged as a significant factor associated with risk of suicidal ideation or attempts (suicidality) in adolescents and young adults. Although the mechanism underlying this association remains uncertain, cognition is a candidate explanatory pathway.

**Method:** We analyzed longitudinal data from the Adolescent Brain Cognitive Development Study (N = 10,228; mean age at end of follow-up 14.17 years, 52.5% male, and 47.5% female). We included three cannabis exposure definitions: one-time experimentation, initiation (more than one-time cannabis use), or ever use. Outcomes included suicidal ideation (SI) and suicide attempt (SA). Mixed-effects regression models quantified associations between cannabis exposures and outcomes adjusting for age, sex, race/ethnicity, household income, and parental education. Additional analyses examined associations of cannabis use with four NIH Toolbox cognitive measures previously linked to suicidality (flanker, working memory, picture vocabulary, and picture sequence memory), and associations of these cognitive measures with SI and SA.

**Results:** Cannabis use was associated with increased odds of SI (ever use odds ratio (OR)=3.18, 95% confidence interval (95%CI) =2.52-4.00) and SA (OR=4.86, 95%CI=3.36-7.03), associations were overall similar across all exposure definitions (all p < 0.001). Cannabis use was also associated with lower performance across multiple cognitive domains (β = −0.08 to −0.16, all p < 0.05). Better cognitive performance, specifically in episodic memory, was associated with lower suicidality: SI (OR = 0.91, 95%CI=0.86-0.95) and SA (OR = 0.88, 95%CI=0.80- 0.97).

**Conclusion:** These findings support cognition as a potential mechanism linking cannabis use to adolescent suicide risk and warrant longitudinal investigation.

## Background

Since 2012, recreational cannabis has been legalized in more than half of U.S states, and its availability continues to expand (1). This policy shift has coincided with higher reports of daily cannabis use and dependence symptoms in young adults (2). Cannabis use is also prevalent in mid and late adolescence, with greater than 30.7% of 12^th^-grade students reporting cannabis use in the last year and over 6% reporting use in the past 30 days (3). As prevalence rises, cannabis use is increasingly recognized as a significant risk factor for suicidal ideation (SI) and attempts (SA) (4). Adolescents who use cannabis carry a 2- to 4-fold elevated risk of SI and SA (5,6), an association that has been replicated across large-scale epidemiologic studies, including inpatient (7,8), and community settings (9). The risk persists into young adulthood and increases in a dose- dependent manner with greater frequency of use (5,6), underlining the public health urgency of understanding the mechanisms through which cannabis elevates suicide risk. Elucidating these mechanisms is a critical step toward developing targeted interventions that address the modifiable pathways through which cannabis use elevates suicide risk.

Cannabis use, particularly when initiated early or used heavily during adolescence, is associated with reduced cognitive functioning across multiple cognitive domains (10,11). The cognitive areas most consistently affected include verbal learning and memory (12,13); executive functions such as working memory, inhibitory control, and cognitive flexibility (12,14); processing speed and attention (12,15); and decision making (12,16). Early onset users show particularly pronounced deficits (17), and adolescent cannabis users perform significantly worse than both alcohol-using and non-using peers on all indices of verbal learning and memory (13).

Independent of cannabis, poor neurocognitive function has been associated with SI and SA in both adult and adolescent populations (18–20). The cognitive domains most consistently implicated include inhibitory control, decision making, and selective attention (19–21). Deficits across both hot (emotional) and cold (non-emotional) executive functioning have been linked to suicidal behavior (22–24). In adolescents specifically, those with a history of SA demonstrate less set-shifting flexibility and greater impulsive decision-making compared to both controls and those with SI (25). Additionally, children with a history of SI are more likely to show deficits in episodic memory (26).

Despite several studies linking cannabis use to cognitive impairment, and cognitive impairment to suicide risk, there are no prior studies evaluating SI, SA, and cognition among cannabis users in adolescence. This study addresses that gap using data from the Adolescent Brain Cognitive Development (ABCD) Study, a nationally representative, longitudinal sample of nearly 12,000 U.S. youth followed from ages 9-10 with yearly assessments that include annual self-report of cannabis use (27), repeated assessment of SI, SA (28), and cognitive testing every two years (29). This design allows prospective examination of how cannabis use in its early stages relates to SI and SA, as well as the cognitive processes that may be associated with cannabis use during adolescence.

The present analysis aimed to: (1) examine whether cannabis use is associated with SI and SA in early adolescence, (2) explore associations between cannabis use and cognitive alterations across multiple domains previously linked to suicidality in the literature, (3) test whether cognitive domains that are associated with cannabis use are independently associated with SI and SA (suicidality) in early adolescence. We hypothesized that cannabis use in early adolescence would be associated with increased suicidality and that cannabis use would be associated with deficits in cognitive domains linked to suicidality.

## Methods and Materials Participants

We included participants from the ABCD Study, which enrolled 11,868 children aged 9 to 10 years at baseline from 21 research sites (17 states) across the United States, recruiting through schools (30,31). The ABCD Study protocol was approved by UC San Diego Institutional Review Board. The current study was exempted from a full review by the Children’s Hospital of Philadelphia IRB. In this study, we used data from ABCD Study release 6.0 including measures collected at baseline (age 9-10), 1-year (age 11), 2-year (age 12), 3-year (age 13), and 4-year (age 14) assessments (30). All participants and parents gave written informed assent and consent, as appropriate, after receiving a complete description of the study.

## Exposure: Cannabis Use

Cannabis use was assessed yearly using youth-report measures, including Low Level Use Questionnaire, Substance Use Interview, Timeline Followback, and mid-year Substance Use Phone Interview (32). For the mid-year phone interview, youth reported substance use in the past 6 months; these responses were integrated into the corresponding subsequent annual assessment wave. Three cannabis use exposure definitions were created to capture distinct use patterns.

Cannabis one-time experimentation was defined as one-time cannabis use (i.e., cannabis puffing or trying), with no evidence of subsequent regular or established use. Cannabis use initiation was defined as reporting more than one puff of cannabis or as regular or established use, based on endorsement of regular use or reported cannabis use across multiple assessment waves (33). Ever cannabis use was defined as meeting the criteria of either experimentation or initiation, reporting the use of any cannabis products, or reporting an age of cannabis use. For descriptive analysis, ever cannabis use was defined as any lifetime cannabis use reported in any of the time points from baseline through 4-year assessment. Participants were coded as missing for the ever-use variable if any contributing time point had missing cannabis-use data, rather than assuming that missing responses indicated no use.

## Outcomes Measures: Suicidal Ideation and Suicide Attempt

Report of SI and SA were assessed using the Kiddie Schedule for Affective Disorders and Schizophrenia for DSM-5 (KSADS-5), a semi-structured diagnostic interview administered to both youth and their parents or guardians (30). For the present study, two binary outcomes were derived: ever SI, indicating any lifetime history of SI reported at any assessment wave, and ever SA, indicating any lifetime history of a SA. Participants with missing data at any contributing wave were coded as missing rather than assumed to be non-endorsers.

## Exploratory Outcome Measure: Cognition

Cognition was assessed with four measures from the ABCD cognitive assessments, which used the NIH Toolbox Cognition Battery (34). We focused our exploratory analysis on 4 tests that assess cognitive traits previously linked to either suicidal ideation or attempts (11,17,19,25,26,35). The Flanker Inhibitory Control and Attention Task (FLNKR) measures response inhibition and selective attention by asking participants to identify a central target stimulus while ignoring flanking distractors. The List Sorting Working Memory Test (LSWMT) assesses working memory capacity by requiring participants to sequence items presented visually and orally in order of size. The Picture Vocabulary Test (PICVCB) measures receptive vocabulary and crystallized intelligence by asking participants to match spoken words to pictures. The Picture Sequence Memory Test (PICSQ) assesses episodic memory by requiring participants to recall and reproduce the sequential order of thematically related pictures.

Uncorrected standard scores were used for all measures. FLNKR, PICVCB, and PICSQ were administered at baseline and at the 2-, 4-year follow-up visits; LSWMT was administered at baseline and at the 4-year follow-up visits only.

## Covariates

All models adjusted for the following covariates selected a priori based on their established associations with cannabis use, suicidality, and cognitive performance. Age was included as a continuous variable. Sex assigned at birth was included as a binary variable (female vs. male). Race/ethnicity was included as a covariate as a proxy for structural racism, racial stress, and related social determinants of health — not as a biological variable or to imply inherent differences between racial or ethnic groups (36). Household income was included as an ordinal variable assessed via a 10-category scale ranging from less than $5,000 to $200,000 or more annually. Parental education was included as an ordinal variable reflecting the highest grade or level of education completed by the most educated parent or guardian in the household, ranging from less than high school to postgraduate degree.

## Statistical analysis

Analyses were conducted using the R version 4.5.1 from March to July 2026. For univariate comparisons, we used t-tests for normally distributed continuous variables, Kruskal-Wallis tests for non-normal continuous variables, and chi-square tests for binary measures. A false discovery rate correction was applied to p-values, with an alpha of 0.05 was used to determine statistical significance.

## Multivariable mixed-effects models

Linear mixed-effects models (which we refer to as “mixed-models” hereafter) were used to estimate associations between cannabis use experimentation, initiation, or ever cannabis use (independent variables), with SI and SA as the dependent variables. These models accounted for the hierarchical and longitudinal nature of the ABCD Study data by including a 4-level hierarchy where survey time points were first nested according to subject ID, then according to family ID, and finally nested according to the research site. We assumed a similar time trend across all subjects and only allowed random intercepts in all mixed models. In the mixed-effects models, all available observations across all assessment waves were included.

## Missing data

We employed listwise deletion by time point, such that a missing value affected only the time point in which it was missing, avoiding participant-level listwise deletion. Rates of missingness are reported for all variables in Table 1.

**Table 1.** Demographic Characteristics by Cannabis Use Status

|  | **Overall** | **No Cannabis** | **Cannabis Use** | **p** | **p (FDR)** | **Missing (%)** |
|---|---|---|---|---|---|---|
| **n** | **10,228** | **9,528** | **700** |  |  |  |
| Age at end of follow-up, mean (SD) | 14.17 (0.71) | 14.14 (0.71) | 14.52 (0.69) | <0.001 | <0.001 | 7.70 |
| Sex, female assigned at birth [N (%)] | 4,856 (47.48) | 4,517 (47.41) | 339 (48.43) | 0.63 | 0.63 | 0.00 |
| **Self-reported race/ethnicity [N (%)]<sup>a</sup>** |  |  |  |  |  |  |
| White | 8,164 (79.82) | 7,651 (80.30) | 513 (73.29) | <0.001 | <0.001 | 0.00 |
| Black | 2,059 (20.13) | 1,861 (19.53) | 198 (28.29) | <0.001 | <0.001 | 0.00 |
| Multiracial | 1,422 (13.90) | 1,302 (13.66) | 120 (17.14) | 0.01 | 0.01 | 0.00 |
| Asian | 730 (7.14) | 696 (7.30) | 34 (4.86) | 0.02 | 0.02 | 0.00 |
| Hawaiian/Pacific Islander | 85 (0.83) | 77 (0.81) | 8 (1.14) | 0.47 | 0.47 | 0.00 |
| AIAN | 493 (4.82) | 433 (4.54) | 60 (8.57) | <0.001 | <0.001 | 0.00 |
| Other | 1,191 (11.64) | 1,075 (11.28) | 116 (16.57) | <0.001 | <0.001 | 0.00 |
| Hispanic | 2,064 (20.18) | 1,893 (19.87) | 171 (24.43) | 0.005 | 0.006 | 0.16 |
| **Household income categories [N (%)]** |  |  |  | <0.001 | <0.001 | 9.55 |
| <\$25,000 | 788 (7.70) | 705 (7.40) | 83 (11.86) |  |  |  |
| \$25,000–50,000 | 945 (9.24) | 857 (8.99) | 88 (12.57) |  |  |  |
| \$50,000–75,000 | 947 (9.26) | 889 (9.33) | 58 (8.29) |  |  |  |
| \$75,000–100,000 | 1,082 (10.58) | 995 (10.44) | 87 (12.43) |  |  |  |
| \$100,000–200,000 | 3,088 (30.19) | 2,946 (30.92) | 142 (20.29) |  |  |  |
| >\$200,000 | 1,609 (15.73) | 1,537 (16.13) | 72 (10.29) |  |  |  |
| **Parent's highest education categories [N (%)]** |  |  |  | <0.001 | <0.001 | 9.64 |
| Less than high school | 375 (3.67) | 335 (3.52) | 40 (5.71) |  |  |  |
| High school graduate | 803 (7.85) | 709 (7.44) | 94 (13.43) |  |  |  |
| Post-high school education | 2,058 (20.12) | 1,875 (19.68) | 183 (26.14) |  |  |  |
| Bachelor's degree | 2,418 (23.64) | 2,290 (24.03) | 128 (18.29) |  |  |  |
| Master's degree or above | 3,588 (35.08) | 3,432 (36.02) | 156 (22.29) |  |  |  |

AIAN = American Indian or Alaska Native; FDR = false discovery rate; SD = standard deviation.
<sup>a</sup> Race categories are not mutually exclusive; participants could endorse multiple races.
FDR correction applied across all comparisons within the table.

## Results

### Sample Characteristics

Participants included 10,228 adolescents (mean age at end of 4-year follow-up 14.17 ± 0.71 years; 48.5% female), of whom 700 (6.8%) reported a history of cannabis use (**Table 1**).

Cannabis users were slightly older than non-users (14.52 vs. 14.14 years, p < 0.001). No sex differences were observed (48.4% vs. 47.4%, p = 0.63). Compared to non-users, cannabis users were more highly represented among youth who self-identified as Black (28.3% vs. 19.5%), American Indian or Alaska Native (8.6% vs. 4.5%), Hispanic (24.4% vs. 19.9%), or multiracial (17.1% vs. 13.7%), and less represented among White (73.3% vs. 80.3%) or Asian youth (4.9% vs. 7.3%; all p < 0.05). Youth reporting cannabis use came from households with lower income (p < 0.001) and were less likely to have a parent with a master’s degree or higher (22.3% vs. 36%) compared to non-users (p < 0.001).

## Cannabis Use and Suicidality

Cannabis users reported higher rates of lifetime suicidality across all outcomes compared to non- users (**Figure 1A, Supplemental Table S1**). SI was more prevalent among cannabis users (47.7% vs. 25.9%, p < 0.001). SA prevalence was more than threefold higher among cannabis users (16.6% vs. 5.2%, p < 0.001). Cannabis users also reported more SI without SA (see **Supplemental Table S1).**

**Figure 1.**
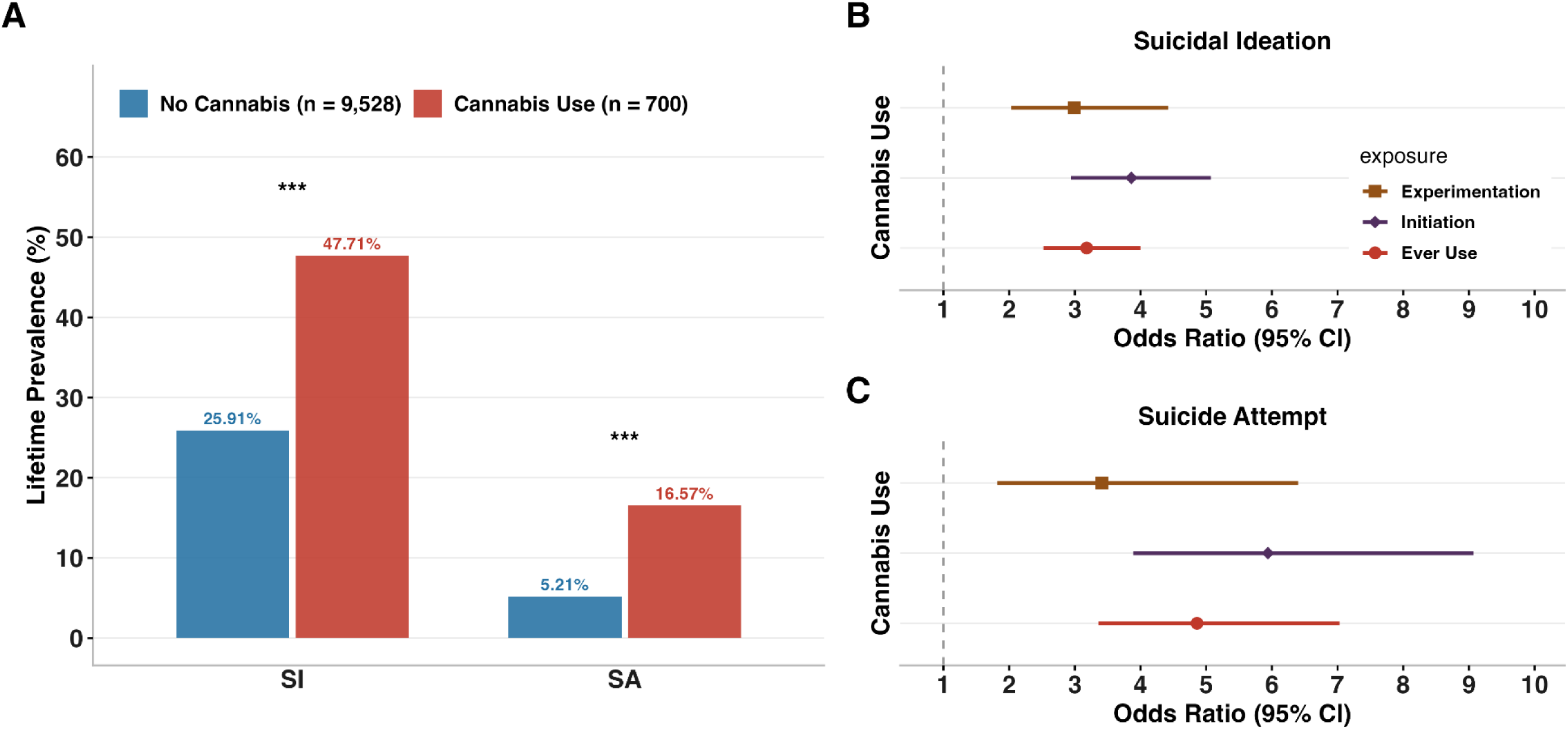
Associations of Cannabis Use with Suicidal Ideation and Attempt A: Lifetime prevalence of suicidal ideation (SI) and suicide attempt (SA) among adolescents reporting any lifetime cannabis use (n = 700) and adolescents reporting no cannabis use (n = 9528). B: Adjusted odds ratios (ORs) and 95% Confidence Interval (CI) for suicidal ideation, estimated in three independent models for each of the 3 cannabis exposure definitions: ever use, experimentation, and initiation. C: Adjusted ORs and 95% CIs for SA, estimated in three independent models for each cannabis exposure. Estimates in panels B and C are derived from mixed-effects logistic regression models adjusted for age, sex assigned at birth, race and ethnicity, household income, and parental education. ***p < 0.001

Multivariable mixed-effects logistic regression models revealed that cannabis use was significantly associated with increased odds of both SI and SA across all three exposure definitions; SA odds ratios were consistently larger than SI (**Figure 1B; Supplemental Table S2-S4**). Cannabis experimentation was associated with increased suicidality (SI: OR = 2.99, 95% CI [2.03–4.42], p < 0.001; SA: OR = 3.41, 95% CI [1.82–6.40], p < 0.001) (**Supplemental Table S2**). Effect sizes were largest in cannabis initiation (SI: OR = 3.86, 95% CI [2.94–5.07], p < 0.001; SA: OR = 5.94, 95% CI [3.86–9.07], p < 0.001) (**Supplemental Table S3**). Ever use of cannabis was associated with three times increased odds of SI (OR = 3.18, 95% CI [2.52– 4.00], p < 0.001) and nearly five times increased odds of SA (OR = 4.86, 95% CI [3.36–7.03], p < 0.001) (**Supplemental Table S4**).

## Cannabis Use and Cognition

Compared to non-users, cannabis users showed lower mean scores across several NIH Toolbox cognitive measures that were previously linked to suicidality (**Figure 2A, Supplemental Table S1**) (26,37,38).

**Figure 2.**
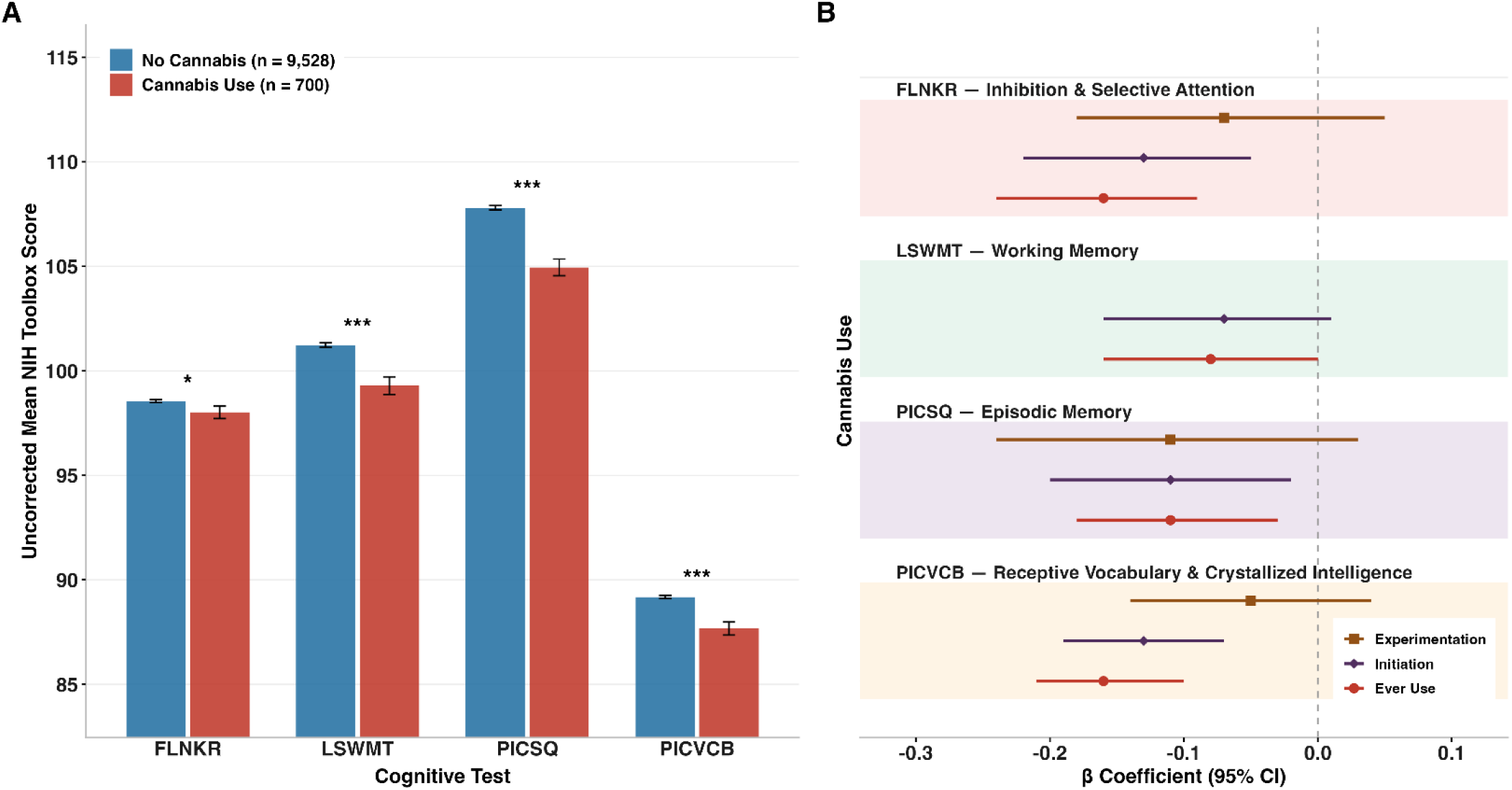
Associations of Cannabis Use with Cognition A: Mean NIH Toolbox scores with 95% Confidence Interval (CI) at the 6-year follow-up across 4 cognitive measures, among adolescents reporting any lifetime cannabis use (n = 700) and adolescents reporting no cannabis use (n = 9,528). B: Adjusted β coefficients and 95% CIs for the association of cannabis use with performance on the Flanker Inhibitory Control and Attention Test, List Sorting Working Memory Test, Picture Vocabulary Test, and Picture Sequence Memory Test, estimated in three independent models for each cannabis exposure: ever use, experimentation, and initiation. Estimates are derived from mixed-effects models using all available observations across assessment waves, adjusted for age, sex assigned at birth, race and ethnicity, household income, and parental education. *p < 0.05; ***p < 0.001 Flanker Inhibitory Control and Attention Task =FLNKR; List Sorting Working Memory Test =LSWMT; Picture Vocabulary Test = PICVCB; Picture Sequence Memory Test =PICSQ.

Differences were statistically significant for working memory (99.29 ± 11.18 vs. 101.23 ± 10.55, p < 0.001), picture sequence memory (104.95 ± 10.60 vs. 107.80 ± 10.45, p < 0.001), and picture vocabulary (87.67 ± 8.30 vs. 89.18 ± 7.71, p < 0.001). The flanker task followed the trend of lower scores in cannabis users but was not statistically significant (98.02 ± 8.01 vs. 98.55 ± 7.00, p = 0.06).

In adjusted multivariable mixed-models, cannabis initiation was significantly associated with deficits in: flanker task (β = −0.13, 95% CI [−0.22, −0.05], p = 0.003), picture sequence memory (β = −0.11, 95% CI [−0.20, −0.02], p = 0.019), and picture vocabulary (β = −0.13, 95% CI [−0.19, −0.07], p < 0.001) but not working memory (β = −0.07, 95% CI [−0.16, 0.01], p = 0.101) (**Figure 2B; Supplemental Table S5**). Associations were similar for ever cannabis use and significant across four domains: flanker task (β = −0.16, 95% CI [−0.24, −0.09], p < 0.001), working memory (β = −0.08, 95% CI [−0.16, −0.00], p =.045), picture sequence memory (β = −0.11, 95% CI [−0.18, −0.03], p =0.007), and picture vocabulary (β = −0.16, 95% CI [−0.21, −0.10], p < 0.001) (**Figure 2B**; **Supplemental Table S6**) which had the largest effect observed across all cognitive outcomes.

In contrast, cannabis experimentation was not significantly associated with any cognitive outcomes (all p >.118, **Figure 2B, Supplemental Table S7**). The mixed-model for cannabis experimentation and working memory could not be estimated because the participant-within- family-within-site random-effects structure resulted in a grouping factor with as many levels as observations. This left no repeated observations within groups, so the model could not estimate the random-effect variance.

## Cognition and Suicidality

Associations between neurocognitive performance and suicidality varied by cognitive domain. Inhibitory control and attention (FLNKR) and working memory (LSWMT) were not significantly associated with SI or SA (all p > .05), though LSWMT showed a trend toward lower SA odds (OR = 0.90, 95% CI [0.80–1.00], p = .057). Episodic memory (PICSQ) was significantly associated with both SI (OR = 0.91, 95% CI [0.86–0.95], p < .001) and SA (OR = 0.88, 95% CI [0.80–0.97], p = .011), such that better memory performance was associated with lower odds of both outcomes (**Figure 3, Supplemental Table S8)**. Receptive vocabulary and crystallized intelligence (PICVCB) was positively associated with SI (OR = 1.10, 95% CI [1.04– 1.17], p = .002), suggesting that higher scores were associated with greater odds of SI, though this measure was not significantly associated with SA (OR = 0.98, 95% CI [0.87–1.11], p = .775).

**Figure 3.**
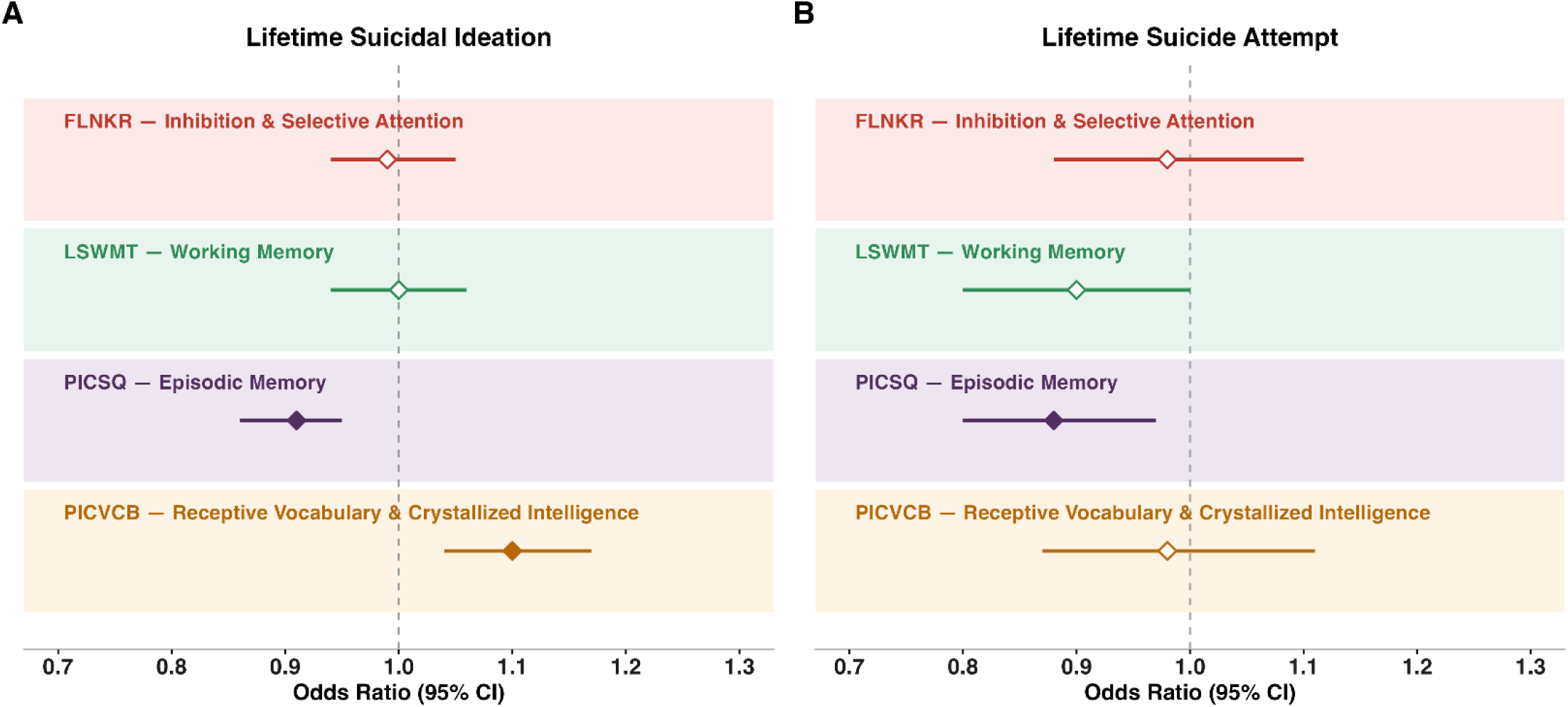
Associations of Cognition with Suicidality Forest plots depict odds ratios (ORs) and 95% confidence intervals from fully adjusted logistic regression models examining associations between four neurocognitive domains and **(A)** lifetime suicidal ideation and **(B)** lifetime suicide attempt. Filled diamonds indicate statistically significant associations (p < .05); open diamonds indicate non-significant associations. The dashed vertical line represents OR = 1.0 (null). Models were adjusted for age, sex, race/ethnicity, household income, and parental education. FLNKR = Flanker Inhibitory Control and Attention Test; LSWMT = List Sorting Working Memory Test; PICSQ = Picture Sequence Memory Test; PICVCB = Picture Vocabulary Test.

## Discussion

In this large, diverse longitudinal cohort, both experimental and repeated cannabis use were independently associated with significantly higher odds of SI and SA after adjustment for demographic and socioeconomic covariates. Notably, even a single reported cannabis exposure was associated with nearly three times the odds of SI and SA, with consistent associations observed for repeated use. Cannabis use was further associated with lower performance across multiple cognitive domains, including inhibitory control, working memory, episodic memory, and receptive vocabulary. Among these, episodic memory was the only domain independently associated with both SI and SA through 4-year follow-up (age 14), consistent with prior ABCD Study work linking suicidality to episodic memory deficits at ages 9–10 (26). Taken together, these findings support a conceptual model in which cannabis-related cognitive impairment may represent one pathway through which cannabis use elevates suicide risk in adolescence (**Figure 4**).

**Figure 4.**
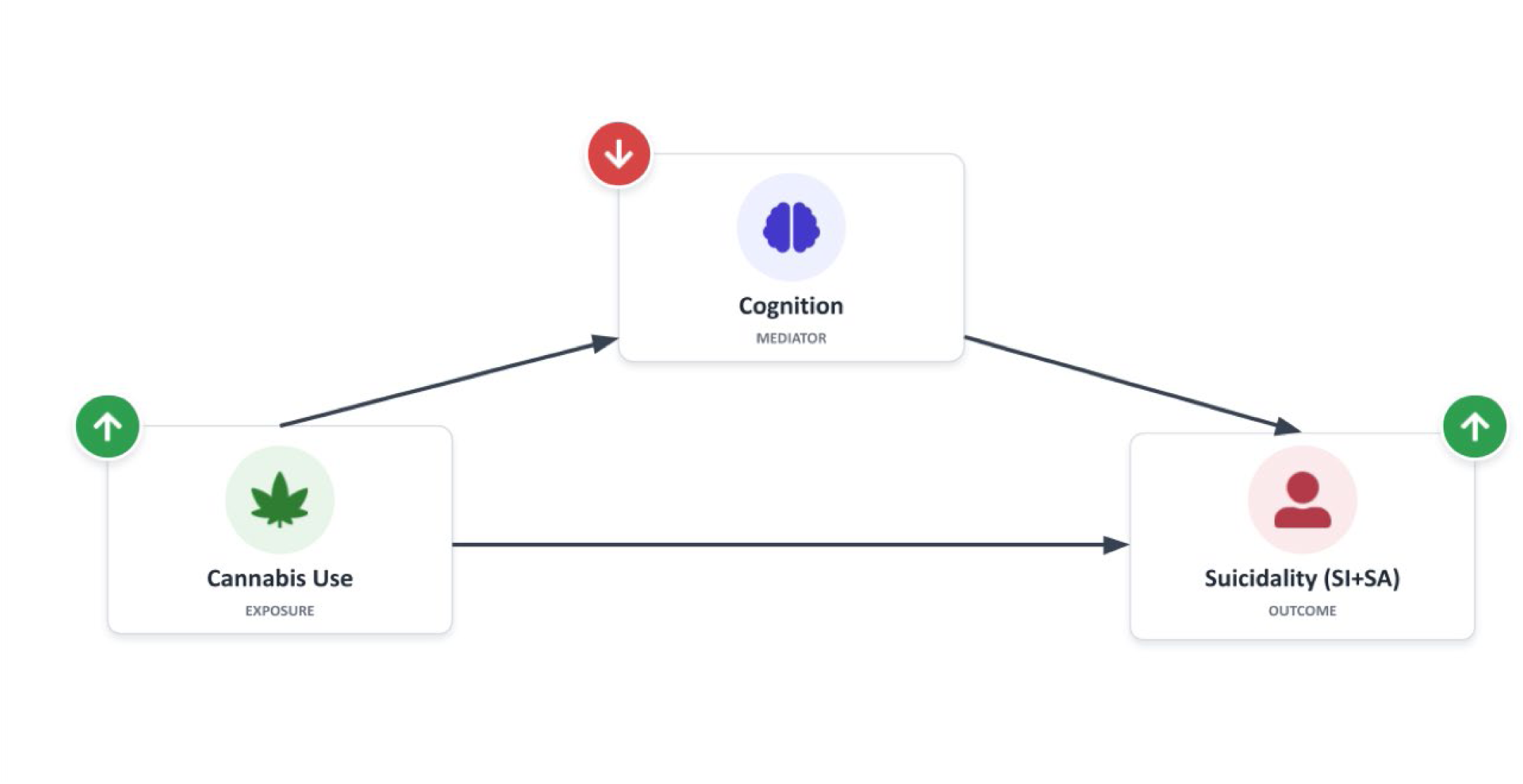
Proposed Model Linking Cannabis Use, Cognition, and Suicidality A conceptual model proposing that neurocognition may partially mediate the association between cannabis use and elevated risk of suicidality. Green arrows indicate positive associations (higher cannabis use associated with greater suicidality); the red arrow indicates a negative association (higher cannabis use associated with lower neurocognitive performance). SI = suicidal ideation; SA = suicide attempt.

By distinguishing one-time cannabis experimentation from initiation, we were able to examine whether suicide risk is confined to regular use and found that it was not. Adolescents reporting only a single cannabis episode showed odds of SI comparable to those with more established use patterns, even after adjustment for demographic and socioeconomic covariates. Although effect sizes were somewhat larger for cannabis initiation, particularly for SA, the confidence intervals overlapped substantially, precluding definitive conclusions about differential risk across exposure levels. Notably, the association between cannabis use and cognitive deficits showed a different pattern than that observed for suicidality: whereas even a single episode of experimentation was associated with elevated suicidality risk, cognitive deficits were only observed among adolescents reporting repeated use. An unexpected finding was the positive association between receptive vocabulary performance and SI. Although this direction of effect has not been widely reported, it is consistent with our prior work in an independent adolescent and young adult cohort, in which participants with SI demonstrated relatively better neurocognitive performance compared to those without (39).

Our study builds on prior ABCD analyses by Wade et al., who used urine and hair toxicology combined with self-report to identify cannabis-using adolescents and found that their cognitive performance fell further behind non-users over time (37). Hair assays, however, detect use only at approximately twice-monthly frequency or greater, and adolescents captured by our experimentation category are precisely those a toxicology-anchored definition would classify as non-users.

Our findings indicate this group is not risk-free: even in the absence of biochemically verified, consistent use, as any level of cannabis exposure was associated with suicidality. This may have clinical implications for pediatric, emergency, and behavioral health settings.

Adolescents indicating any affirmative response to a cannabis screening question may warrant consideration of a structured suicide risk assessment, rather than reserving deeper evaluation for adolescents endorsing regular or problematic use. A brief cannabis screener could therefore function as a sentinel marker for elevated suicide risk that would otherwise be missed.

A key innovation of this work is that it ties together cannabis use, suicidality outcomes, and cognitive alterations. We propose a mechanistic model in which adolescent cannabis use may contribute to suicide risk partly through its effects on cognitive processing. Adolescence is a period of protracted maturation of prefrontal circuits that support inhibition, working memory, and decision-making, processes underpinned by an endocannabinoid system that exogenous cannabinoids can disrupt (10,40). Deficits in these domains have been repeatedly implicated in suicidality (20,25). Functional neuroimaging studies have shown aberrant activation patterns in prefrontal, anterior cingulate, and limbic circuits during tasks requiring working memory, response inhibition, executive control, reward processing, and emotional processing in chronic adolescent cannabis users (41,42). Consistent with this model, adolescent-onset cannabis use has been linked to neuropsychological decline persisting into adulthood (38), with particularly pronounced deficits among early-onset users (17) and trajectories that diverge and widen across adolescence (37). However, these cannabis-related cognitive deficits are diminished during periods of abstinence (43). Because cannabis use became prevalent only in the most recent ABCD assessment wave, the available data do not yet provide the temporal separation required to directly evaluate the proposed longitudinal model, nor did the current study examine neuroimaging correlates of the observed cognitive deficits. Continued follow-up of the cohort, with integration of structural and functional neuroimaging data, will provide an important opportunity to prospectively test the hypothesized temporal and neurobiological associations as additional post-exposure outcome data accrue.

Several limitations deserve consideration. First, cannabis use was only classified by self- report in this study, which is subject to recall error and underreporting; misclassification of this kind attenuate or inflate the associations reported. Nevertheless, we still observe robust associations between cannabis use and SI and SA, consistent with other studies (5,7,8). Second, the current analysis is constrained by the availability of cognitive data in the latest ABCD release, as data collection is ongoing. Future releases will provide richer longitudinal cognitive phenotyping during developmental periods marked by increasing rates of cannabis use and suicidality. Such data will be critical for refining and extending the present findings. Third, our analyses do not infer causation, and they do not constitute a formal test of mediation.

Forthcoming ABCD data releases extending follow-up into young adulthood will provide sufficient temporal separation and support longitudinal mediation to formally test our proposed pathway linking cannabis, cognition, and suicidality.

In conclusion, we report elevated suicidality risk among early adolescent cannabis users that was not confined to established, regular use. Adolescents reporting only a single episode of experimentation carried increased odds of suicidality comparable to those with initiated, regular use. The consistency of these associations across exposure levels, combined with independent links between cannabis use and cognitive deficits and between episodic memory and suicidality, provides support for a model in which cognitive impairment may partially mediate the pathway from cannabis use to suicidality (26,34,35). Future analyses of longitudinal cohorts such as ABCD will be important for testing the temporal and mechanistic relationships underlying this proposed pathway as participants age into young adulthood.

## Data Availability

All data produced in the present study are available upon reasonable request to the authors

## Notes

### Competing Interest Statement

The authors have declared no competing interest.

### Author Declarations

IRB of Childrens Hospital of Philadelphia gave ethical approval for this work

### Funding

- National Institute of Mental Health: R25MH119043, R01MH134886, P50MH115838
- American Foundation for Suicide Prevention: SRG-0-006-22, ECR-0-197-25

