## Supplemental Table S1 for "Cannabis Use, Cognition, and Suicide Risk in Adolescents from the Adolescent Brain Cognitive Development Study"

**Supplemental Material**

**Supplemental Table S1.** Mental Health and Cognitive Outcomes by Cannabis Use Status
**Supplemental Table S2.** Association of Cannabis Use Experimentation with Suicidal Ideation and Suicide Attempt
**Supplemental Table S3.** Association of Cannabis Use Initiation with Suicidal Ideation and Suicide Attempt
**Supplemental Table S4.** Association of Ever Cannabis Use with Suicidal Ideation and Suicide Attempt
**Supplemental Table S5.** Association of Cannabis Use Initiation with Cognitive Performance
**Supplemental Table S6.** Association of Ever Cannabis Use with Cognitive Performance
**Supplemental Table S7.** Association of Cannabis Use Experimentation with Cognitive Performance
**Supplemental Table S8.** Association of Cognitive Performance and Suicidality

**Supplemental Table S1. Mental Health and Cognitive Outcomes by Cannabis Use Status**

| | Overall | No Cannabis | Cannabis Use | p | p (FDR) | Missing (%) |
|---|---|---|---|---|---|---|
| **n** | 10,228 | 9,528 | 700 | | | |
| **Suicidality [N(%)]** | | | | | | |
| SI | 2,803 (27.41) | 2,469 (25.91) | 334 (47.71) | <0.001 | <0.001 | 14.81 |
| SA | 612 (5.98) | 496 (5.21) | 116 (16.57) | <0.001 | <0.001 | 18.70 |
| SI without SA | 2,548 (24.91) | 2,260 (23.72) | 288 (41.14) | <0.001 | <0.001 | 15.48 |
| **NIH Toolbox Score [Uncorrected Mean Score, SD]** | | | | | | |
| FLNKR | 98.52 (7.07) | 98.55 (7.00) | 98.02 (8.01) | 0.05 | 0.06 | 0.12 |
| LSWMT | 101.10 (10.60) | 101.23 (10.55) | 99.29 (11.18) | <0.001 | <0.001 | 0.25 |
| PICSQ | 107.61 (10.49) | 107.80 (10.45) | 104.95 (10.60) | <0.001 | <0.001 | 0.03 |
| PICVCB | 89.07 (7.76) | 89.18 (7.71) | 87.67 (8.30) | <0.001 | <0.001 | 0.03 |

**Note.** Values shown as mean (SD) or N (%). Missing (%) = % with missing data. p-values from chi-square tests or t-tests; FDR = Benjamini-Hochberg. SA = suicide attempt; SI = suicidal ideation, FLNKR = Flanker Inhibitory Control and Attention Test; LSWMT = List Sorting Working Memory Test; PICSQ = Picture Sequence Memory Test; PICVCB = Picture Vocabulary Test. NIH Toolbox scores uncorrected, average from baseline to wave-4.

**Supplemental Table S2.** Association of Cannabis Use Experimentation with Suicidal Ideation and Suicide Attempt

| | SI OR (95% CI) | p-value | SA OR (95% CI) | p-value |
|---|---|---|---|---|
| | n=10, 473 | | n=10, 473 | |
| **Cannabis Experimentation (One-time Use)** | **2.99 [2.03–4.42]** | **<0.001** | **3.41 [1.82–6.40]** | **<0.001** |
| Age | 1.23 [1.19–1.28] | **<0.001** | 1.53 [1.41–1.65] | **<0.001** |
| Sex | 1.73 [1.54–1.95] | **<0.001** | 1.75 [1.36–2.26] | **<0.001** |
| Black | 1.32 [1.07–1.62] | **0.008** | 1.51 [0.99–2.28] | 0.053 |
| White | 1.30 [1.06–1.59] | **0.012** | 1.31 [0.85–2.00] | 0.220 |
| Hispanic | 1.14 [0.96–1.35] | 0.137 | 0.98 [0.68–1.41] | 0.918 |
| Household income | 0.94 [0.91–0.97] | **<0.001** | 0.90 [0.84–0.97] | **0.003** |
| Parental education | 1.03 [0.99–1.07] | 0.128 | 0.99 [0.91–1.07] | 0.712 |

**Note.** All ORs, 95% CIs, and p-values from a fully adjusted mixed-effects logistic regression with random effects for participant, family, and site. Suicide attempt = SA; suicidal ideation =SI; OR= odds ratio.

**Supplemental Table S3.** Association of Cannabis Use Initiation with Suicidal Ideation and Suicide Attempt

| | SI OR (95% CI) | p-value | SA OR (95% CI) | p-value |
|---|---|---|---|---|
| | n=11, 196 | | n=11,196 | |
| **Cannabis Initiation (Regular Use)** | **3.86 [2.94–5.07]** | <0.001 | **5.94 [3.89–9.07]** | <0.001 |
| Age | 1.15 [1.12–1.18] | **<0.001** | 1.36 [1.28–1.44] | **<0.001** |
| Sex | 1.53 [1.38–1.69] | **<0.001** | 1.64 [1.31–2.04] | **<0.001** |
| Black | 1.24 [1.04–1.48] | **0.019** | 1.37 [0.95–1.97] | 0.087 |
| White | 1.14 [0.96–1.36] | 0.137 | 1.05 [0.73–1.50] | 0.800 |
| Hispanic | 1.07 [0.92–1.25] | 0.380 | 1.10 [0.81–1.50] | 0.547 |
| Household income | 0.95 [0.92–0.98] | **<0.001** | 0.91 [0.86–0.97] | **0.002** |
| Parental education | 1.03 [0.99–1.06] | 0.139 | 0.99 [0.93–1.06] | 0.806 |

**Note.** All ORs, 95% CIs, and p-values from a fully adjusted mixed-effects logistic regression with random effects for participant, family, and site. Suicide attempt = SA; suicidal ideation =SI; OR= odds ratio.

**Supplemental Table S4.** Association of Ever Cannabis Use with Suicidal Ideation and Suicide Attempt

| | SI OR (95% CI) | p-value | SA OR (95% CI) | p-value |
|---|---|---|---|---|
| | n=11, 570 | | n=11, 570 | |
| **Cannabis Ever Use (Any Lifetime Use)** | **3.18 [2.52–4.00]** | **<0.001** | **4.86 [3.36–7.03]** | **<0.001** |
| Age | 1.15 [1.12–1.17] | **<0.001** | 1.35 [1.28–1.42] | **<0.001** |
| Sex | 1.41 [1.28–1.56] | **<0.001** | 1.48 [1.20–1.82] | **<0.001** |
| Black | 1.26 [1.06–1.49] | **0.009** | 1.40 [1.00–1.97] | 0.051 |
| White | 1.17 [0.99–1.38] | 0.066 | 1.11 [0.79–1.55] | 0.558 |
| Hispanic | 1.06 [0.92–1.23] | 0.409 | 1.06 [0.79–1.41] | 0.716 |
| Household income | 0.96 [0.94–0.99] | **0.004** | 0.91 [0.87–0.96] | **<0.001** |
| Parental education | 1.02 [0.99–1.06] | 0.153 | 0.99 [0.93–1.06] | 0.807 |

**Note.** All ORs, 95% CIs, and p-values from a fully adjusted mixed-effects logistic regression with random effects for participant, family, and site. Suicide attempt = SA; suicidal ideation =SI; OR= odds ratio.

**Supplemental Table S5.** Association of Cannabis Use Initiation with Cognitive Performance

| | FLNKR β (95% CI) | p | LSWMT β (95% CI) | p | PICSQ β (95% CI) | p | PICVCB β (95% CI) | p |
|---|---|---|---|---|---|---|---|---|
| | n=10,244 | | n=9,928 | | n=10,739 | | n=10,750 | |
| **Cannabis Initiation** | **-0.13 [-0.22 – -0.05]** | **0.003** | -0.07 [-0.16 – 0.01] | 0.101 | **-0.11 [-0.20 – -0.02]** | **0.019** | **-0.13 [-0.19 – -0.07]** | <0.001 |
| Age | 0.23 [0.23 – 0.24] | **<0.001** | 0.15 [0.14 – 0.16] | **<0.001** | 0.14 [0.13 – 0.14] | **<0.001** | 0.22 [0.22 – 0.23] | **<0.001** |
| Sex | -0.06 [-0.09 – -0.03] | **<0.001** | -0.03 [-0.06 – 0.00] | 0.052 | 0.12 [0.09 – 0.15] | **<0.001** | -0.04 [-0.06 – -0.01] | **0.007** |
| Black | -0.17 [-0.22 – -0.12] | **<0.001** | -0.21 [-0.27 – -0.16] | **<0.001** | -0.29 [-0.34 – -0.23] | **<0.001** | -0.35 [-0.40 – -0.30] | **<0.001** |
| White | 0.09 [0.04 – 0.14] | **<0.001** | 0.17 [0.11 – 0.22] | **<0.001** | 0.12 [0.06 – 0.17] | **<0.001** | 0.25 [0.20 – 0.30] | **<0.001** |
| Hispanic | -0.03 [-0.08 – 0.01] | 0.113 | -0.08 [-0.13 – -0.04] | **<0.001** | -0.02 [-0.07 – 0.02] | 0.321 | -0.28 [-0.32 – -0.24] | **<0.001** |
| Household income | 0.03 [0.03 – 0.04] | **<0.001** | 0.05 [0.04 – 0.06] | **<0.001** | 0.03 [0.02 – 0.04] | **<0.001** | 0.04 [0.04 – 0.05] | **<0.001** |
| Parental education | 0.04 [0.03 – 0.05] | **<0.001** | 0.06 [0.05 – 0.07] | **<0.001** | 0.04 [0.03 – 0.05] | **<0.001** | 0.09 [0.09 – 0.10] | **<0.001** |

**Note.** β = Standardized regression coefficient; CI = Confidence Interval. FLNKR = Flanker Inhibitory Control and Attention Test; LSWMT = List Sorting Working Memory Test; PICSQ = Picture Sequence Memory Test; PICVCB = Picture Vocabulary Test. Models are fully adjusted and include age, sex, race/ethnicity, household income, and parental education as covariates.

**Supplemental Table S6.** Association of Ever Cannabis Use with Cognitive Performance

| | FLNKR β (95% CI) | p | LSWMT β (95% CI) | p | PICSQ β (95% CI) | p | PICVCB β (95% CI) | p |
|---|---|---|---|---|---|---|---|---|
| | n=11,321 | | n= 11,165 | | n=11,399 | | n=11,404 | |
| **Cannabis Ever Use** | **-0.16 [-0.24 – -0.09]** | **<0.001** | **-0.08 [-0.16 – -0.00]** | **0.045** | **-0.11 [-0.18 – -0.03]** | **0.007** | **-0.16 [-0.21 – -0.10]** | **<0.001** |
| Age | 0.25 [0.24 – 0.25] | **<0.001** | 0.16 [0.15 – 0.16] | **<0.001** | 0.14 [0.14 – 0.15] | **<0.001** | 0.23 [0.23 – 0.23] | **<0.001** |
| Sex | -0.06 [-0.08 – -0.03] | **<0.001** | -0.05 [-0.07 – -0.02] | **0.002** | 0.11 [0.08 – 0.13] | **<0.001** | -0.04 [-0.06 – -0.01] | **0.002** |
| Black | -0.16 [-0.21 – -0.12] | **<0.001** | -0.20 [-0.26 – -0.15] | **<0.001** | -0.29 [-0.34 – -0.24] | **<0.001** | -0.32 [-0.37 – -0.28] | **<0.001** |
| White | 0.09 [0.05 – 0.14] | **<0.001** | 0.16 [0.11 – 0.21] | **<0.001** | 0.11 [0.06 – 0.16] | **<0.001** | 0.26 [0.21 – 0.30] | **<0.001** |
| Hispanic | -0.03 [-0.07 – 0.01] | 0.180 | -0.08 [-0.12 – -0.04] | **<0.001** | -0.02 [-0.06 – 0.02] | 0.439 | -0.27 [-0.31 – -0.23] | **<0.001** |
| Household income | 0.04 [0.03 – 0.04] | **<0.001** | 0.05 [0.04 – 0.06] | **<0.001** | 0.03 [0.02 – 0.04] | **<0.001** | 0.04 [0.04 – 0.05] | **<0.001** |
| Parental education | 0.04 [0.03 – 0.05] | **<0.001** | 0.07 [0.06 – 0.08] | **<0.001** | 0.04 [0.03 – 0.05] | **<0.001** | 0.09 [0.08 – 0.09] | **<0.001** |

**Note.** β = Standardized regression coefficient; CI = Confidence Interval. FLNKR = Flanker Inhibitory Control and Attention Test; LSWMT = List Sorting Working Memory Test; PICSQ = Picture Sequence Memory Test; PICVCB = Picture Vocabulary Test. Models are fully adjusted and include age, sex, race/ethnicity, household income, and parental education as covariates.

**Supplemental Table S7.** Association of Cannabis Use Experimentation with Cognitive Performance

| | FLNKR β (95% CI) | p | LSWMT β (95% CI) | p | PICSQ β (95% CI) | p | PICVCB β (95% CI) | p |
|---|---|---|---|---|---|---|---|---|
| | n=8,210 | | | | n=9,347 | | n=9,359 | |
| **Cannabis Experimentation** | -0.07 [-0.18 – 0.05] | 0.254 | — | — | -0.11 [-0.24 – 0.03] | 0.118 | -0.05 [-0.14 – 0.04] | 0.282 |
| Age | 0.17 [0.16 – 0.18] | **<0.001** | — | — | 0.09 [0.08 – 0.11] | **<0.001** | 0.23 [0.22 – 0.24] | **<0.001** |
| Sex | -0.09 [-0.12 – -0.05] | **<0.001** | — | — | 0.12 [0.08 – 0.15] | **<0.001** | -0.04 [-0.07 – -0.01] | **0.009** |
| Black | -0.18 [-0.24 – -0.12] | **<0.001** | — | — | -0.28 [-0.34 – -0.21] | **<0.001** | -0.36 [-0.41 – -0.30] | **<0.001** |
| White | 0.08 [0.02 – 0.13] | **0.007** | — | — | 0.13 [0.07 – 0.19] | **<0.001** | 0.23 [0.17 – 0.28] | **<0.001** |
| Hispanic | -0.06 [-0.11 – -0.01] | **0.017** | — | — | -0.04 [-0.09 – 0.02] | 0.163 | -0.28 [-0.33 – -0.24] | **<0.001** |
| Household income | 0.04 [0.03 – 0.05] | **<0.001** | — | — | 0.03 [0.02 – 0.04] | **<0.001** | 0.05 [0.04 – 0.06] | **<0.001** |
| Parental education | 0.04 [0.03 – 0.05] | **<0.001** | — | — | 0.04 [0.03 – 0.05] | **<0.001** | 0.10 [0.09 – 0.11] | **<0.001** |

**Note.** β = Standardized regression coefficient; CI = Confidence Interval. FLNKR = Flanker Inhibitory Control and Attention Test; LSWMT = List Sorting Working Memory Test; PICSQ = Picture Sequence Memory Test; PICVCB = Picture Vocabulary Test. Models are fully adjusted and include age, sex, race/ethnicity, household income, and parental education as covariates. LSWMT × Experimentation model was not available in the data. Bold p-values indicate statistical significance ($p < 0.05$). — indicates data not available.

**Supplemental Table S8.** Association of Cognitive Performance and Suicidality

| Predictor | SI OR (95% CI) | p-value | SA OR (95% CI) | p-value |
|---|---|---|---|---|
| | n= 11,284 | | n=11,283 | |
| **FLNKR** | 0.99 [0.94–1.05] | 0.771 | 0.98 [0.88–1.10] | 0.741 |
| Age | 1.13 [1.10–1.17] | **<0.001** | 1.33 [1.25–1.41] | **<0.001** |
| Sex | 1.33 [1.21–1.48] | **<0.001** | 1.47 [1.20–1.80] | **<0.001** |
| Black | 1.18 [0.99–1.41] | 0.071 | 1.27 [0.91–1.77] | 0.164 |
| White | 1.17 [0.98–1.40] | 0.090 | 1.06 [0.76–1.48] | 0.735 |
| Hispanic | 1.00 [0.86–1.16] | 0.992 | 1.11 [0.84–1.47] | 0.471 |
| Household income | 0.96 [0.94–0.99] | **0.020** | 0.91 [0.86–0.96] | **<0.001** |
| Parental education | 1.02 [0.98–1.06] | 0.272 | 0.98 [0.92–1.04] | 0.503 |
| | n=11,102 | | n=11,101 | |
| **LSWMT** | 1.00 [0.94–1.06] | 0.996 | 0.90 [0.80–1.00] | 0.057 |
| Age | 1.13 [1.10–1.15] | **<0.001** | 1.31 [1.25–1.38] | **<0.001** |
| Sex | 1.37 [1.23–1.52] | **<0.001** | 1.57 [1.28–1.93] | **<0.001** |
| Black | 1.14 [0.96–1.37] | 0.140 | 1.13 [0.81–1.59] | 0.471 |
| White | 1.13 [0.95–1.36] | 0.171 | 1.02 [0.73–1.42] | 0.925 |
| Hispanic | 1.01 [0.87–1.18] | 0.849 | 1.21 [0.91–1.59] | 0.185 |
| Household income | 0.96 [0.93–0.99] | **0.013** | 0.89 [0.85–0.95] | **<0.001** |
| Parental education | 1.03 [0.99–1.06] | 0.148 | 1.02 [0.96–1.10] | 0.480 |
| | n=11,376 | | n=11,376 | |
| **PICSQ** | **0.91 [0.86–0.95]** | **<0.001** | **0.88 [0.80–0.97]** | **0.011** |
| Age | 1.15 [1.12–1.18] | **<0.001** | 1.35 [1.28–1.42] | **<0.001** |
| Sex | 1.40 [1.27–1.55] | **<0.001** | 1.53 [1.26–1.86] | **<0.001** |
| Black | 1.15 [0.97–1.37] | 0.112 | 1.16 [0.84–1.59] | 0.376 |
| White | 1.15 [0.96–1.36] | 0.121 | 1.01 [0.74–1.39] | 0.946 |
| Hispanic | 1.04 [0.90–1.20] | 0.591 | 1.10 [0.85–1.44] | 0.466 |
| Household income | 0.96 [0.93–0.99] | **0.005** | 0.90 [0.86–0.95] | **<0.001** |
| Parental education | 1.04 [1.00–1.07] | **0.031** | 1.01 [0.95–1.07] | 0.826 |
| Table continues below | | | | |

| Predictor | SI OR (95% CI) | p-value | SA OR (95% CI) | p-value |
|---|---|---|---|---|
| | n=11,381 | | n=11,381 | |
| **PICVCB** | **1.10 [1.04–1.17]** | **0.002** | 0.98 [0.87–1.11] | 0.775 |
| Age | 1.11 [1.08–1.14] | **<0.001** | 1.33 [1.26–1.41] | **<0.001** |
| Sex | 1.40 [1.27–1.55] | **<0.001** | 1.51 [1.24–1.83] | **<0.001** |
| Black | 1.22 [1.02–1.45] | **0.026** | 1.20 [0.87–1.65] | 0.266 |
| White | 1.10 [0.93–1.31] | 0.255 | 1.01 [0.74–1.39] | 0.929 |
| Hispanic | 1.06 [0.92–1.23] | 0.426 | 1.11 [0.85–1.44] | 0.440 |
| Household income | 0.95 [0.93–0.98] | **<0.001** | 0.90 [0.85–0.95] | **<0.001** |
| Parental education | 1.02 [0.99–1.06] | 0.234 | 1.01 [0.95–1.07] | 0.821 |

**Note.** All β coefficients, 95% CIs, and p-values from a fully adjusted mixed-effects linear regression with random effects for participant, family, and site. Outcomes are NIH Toolbox uncorrected scores (wave 4). FLNKR= Flanker Inhibitory Control and Attention Task; LSWMT= List Sorting Working Memory Test; PICVCB =Picture Vocabulary Test ; PICSQ = Picture Sequence Memory Test; SA= Suicide attempt; SI= suicidal ideation; OR= odds ratio
